# Long-read sequencing resolves functional CYP2D6 alleles tagged by a cocaine use disorder–associated haplotype

**DOI:** 10.64898/2026.11.23.26368775

**Authors:** Michael R. Garvin, Daniel M. Rosenblum, Andrew W. Bergen, Stanley H. Weiss

**Affiliations:** Oregon Research Institute, Springfield, OR; Williwaw Biosciences, LLC, Clarkston, MI; Department of Medicine, Rutgers New Jersey Medical School, Newark NJ

**Keywords:** long-read sequencing, copy number variation, addiction, cocaine, CYP2D6

## Abstract

The CYP2D6 locus exhibits extensive allelic diversity and structural complexity, complicating the identification of functional variants underlying genetic associations. We previously identified a four-single-nucleotide polymorphism (SNP) haplotype (*AACG*) associated with cocaine use disorder and altered *CYP2D6* expression in the brain. To characterize the molecular variation of this haplotype, we performed targeted PacBio long-read sequencing on 19 individuals of European ancestry carrying zero, one, or two copies of *AACG*. Complementary analyses included human pangenome assemblies, protein structural modeling, splice quantitative trait loci, and SNP-array probe sequences. Long-read sequencing resolved distinct *CYP2D6* alleles associated with *AACG*, including *CYP2D6* star *2* and star *4*. Four of five individuals lacking *AACG* carried two canonical reference alleles, whereas nearly all *AACG* homozygotes carried noncanonical alleles. The star 2 allele contained two missense substitutions (R296C and S486T), while star 4 carried four missense substitutions (P34S, L91M, H94R, and S486T) and a loss-of-function splice-site mutation. Structural comparisons suggested potential substrate-dependent effects on ligand binding, while publicly available transcriptomic data associated *AACG* variants with altered splice-junction usage across multiple tissues, including brain. Although SNP-array intensity differences initially suggested copy-number variation, long-read sequencing did not support consistent gene dosage differences. Instead, polymorphisms within array probe-binding regions provided a plausible explanation for these signals. These findings demonstrate that a disease-associated SNP haplotype tags functionally distinct *CYP2D6* alleles and highlight the importance of haplotype-resolved sequencing for interpreting genetic associations at complex pharmacogenomic loci.

## INTRODUCTION

Cytochrome P450 enzymes play a central role in the metabolism of pharmaceutical drugs, endogenous compounds, and environmental xenobiotics (1). Cytochrome P450 family 2 subfamily D member 6 (CYP2D6) is particularly important because it metabolizes approximately one-quarter of clinically used medications, including many prescribed for psychiatric conditions (2). Genetic variation in *CYP2D6* is a well-established determinant of drug metabolism and treatment response, but its potential contribution to susceptibility to psychiatric and substance use disorders remains less well understood (3).

We previously applied a novel approach to investigate the contribution of epistasis to the high heritability of cocaine use disorder (CUD). This analysis identified a four-single-nucleotide polymorphism (SNP) haplotype within *CYP2D6* that was strongly associated with cocaine use in individuals of European ancestry and was linked to altered *CYP2D6* expression in the brain (4). However, the *CYP2D6* locus is structurally complex and can exhibit extensive allelic diversity, copy-number variation (CNV), as well as high sequence similarity to nearby pseudogenes. These features complicate short-read sequencing and array-based genotyping and limit the ability to identify the functional variants underlying trait associations.

To investigate the genomic features linked to this haplotype, we initially analyzed long-read assemblies from the Human Pangenome Project (5). These analyses revealed multiple potentially causal variants associated with the risk haplotype, including four missense substitutions in the CYP2D6 protein, a splice site alteration adjacent to exon 4, and the loss of an interferon-gamma response element downstream of the gene. Although some assemblies also exhibited variation in *CYP2D6* copy number, CNV was not consistently associated with the risk haplotype. Nevertheless, differences in raw SNP-array signal intensity correlated with the number of risk-haplotype copies, initially suggesting that the association might reflect underlying structural variation (4).

Because the pangenome assemblies were derived from individuals with unknown cocaine use status, we next sought to establish whether these sequence variants were present in individuals from our cocaine use cohort. To that end, we performed targeted PacBio long-read sequencing of the *CYP2D6* locus in 19 individuals with 0, 1, or 2 copies of the eQTL risk allele. These data confirmed that the risk allele is linked to a phased haplotype carrying five functional alterations (P34S, L91M, H94R, the splice site mutation c.506-G.A, and S486T), which corresponds to the pharmacogenomically defined *CYP2D6*4 allele.* A second linked haplotype observed in individuals with one or two copies of the risk allele carried R296C and S486T, consistent with *CYP2D6*\*2. By contrast, four of five individuals lacking the risk allele carried two copies of the *CYP2D6* gene identical to the reference sequence. We also investigated the basis of the SNP-array intensity differences and identified probe-sequence mismatches involving neighboring polymorphisms as an alternative explanation to CNV. Together, these findings demonstrate that the cocaine use-associated haplotype distinguishes *CYP2D6* alleles with different functional properties, providing a molecular framework for investigating the contribution of CYP2D6 variation to cocaine susceptibility.

## METHODS

### Samples

Nineteen samples of European ancestry were chosen that carried 0, 1, or 2 copies of the *AACG* risk allele, different levels of cocaine use, and varied in LogRR intensity values for the SNPs within the body of the *CYP2D6* gene (Table S1).

### Sequencing and bioinformatics

Sequencing was performed by Eremid Genomic Services. High molecular weight genomic DNA (gDNA) was extracted from approximately 3 million cells using the Nanobind^TM^ CBB kit (PacBio^®^). gDNA was processed through the Twist-PacBio workflow for Long Read Library Preparation and Standard Hyb v2 Enrichment (Twist^®^) in conjunction with the Twist Alliance Long-Read PGx Panel (Table S2). In brief, 2 µg of each sample were fragmented using Covaris^®^ g-Tubes to 10 Kb fragments size. The gDNA fragments were end repaired, ligated to Twist^®^ Universal Adapters, and amplified using UDI barcoded primers and the KOD Xtreme Hot Start DNA Polymerase (MilliporeSigma^®^). The resulting libraries were combined at equal amounts in three pools of 6 or 7 samples. 4 µg of each pool was hybridized to the Twist^®^ Alliance Long-Read PGx panel at 70°C for 16 hours, bead purified, and amplified for 18 cycles. The three pools of enriched libraries were combined at equal amounts and 900 ng were end repaired and ligated to SMRTbell^®^ Adapters using the SMRTbell^®^ prep kit 3.0 (PacBio^®^). The final library was loaded into one SMRTcell-25M and ran on the Revio system with a movie duration of 30 hours.

HiFi reads provided by Eremid were imported into CLC Genomics Workbench version 26.0. The mean read length across samples was 3,290 bp, with sample means ranging from 2,975 to 3,586 bp (Table S3). Although the mean read length was shorter than the 4,218-bp *CYP2D6* gene, overlapping reads provided sufficient coverage to assemble contigs spanning the coding region and the variants used for haplotype assignment. To extract *CYP2D6*-specific captured long read sequences for further analysis, the entire bam file for each individual was mapped to the Twist PGx panel reference used by Eremid to capture fragments initially using default parameters.

The mapped sequences from each individual were then assembled independently using the De Novo Assembly tool in CLC Genomics. Automatic word-size selection was enabled, the minimum reported contig length was 3000 bp, and reads were mapped back to the resulting contigs with consensus updating enabled. Read-to-contig mapping used mismatch, insertion, and deletion costs of 2, 3, and 3, respectively. Initially, the length fraction and similarity fraction were set to 0.95. If fully assembled alleles were not obtained with these settings, the length fraction was reduced to 0.9, 0.8, 0.7 and finally 0.6 until both alleles were assembled.

Assignment to the *CYP2D6* gene was based on multiple paralog-discriminating positions distributed across the locus, including the diagnostic indel at position 138 distinguishing *CYP2D7* and *CYP2D6*. Alignments were also inspected for regional changes in paralog identity indicative of *CYP2D6–CYP2D7* hybrid alleles. After each allele was called, the assembly for the given contig was queried to identify any heterozygous sites, indicating an additional allele. Finally, to verify each called allele, all *CYP2D6*-extracted reads from the original Eremid derived bam file were mapped to the *CYP2D6* locus from the human genome hg38 assembly which also included 5kb of flanking sequence to allow for mapping of any overlapping fragment(s). These assemblies were inspected to verify called alleles and any heterozygous sites that might indicate two nearly identical alleles (if an individual harbored two completely identical alleles, then they would of course not be identified).

For structural analyses of the CYP2D6 protein, experimentally determined protein structures were obtained from the Protein Data Bank (PDB IDs: 4WNW and 5TFT) and examined using UCSF Chimera (5,6). Structural visualization and geometric measurements were performed using Chimera’s Structure Analysis tools. Interatomic distances were measured using the Distances tool, while bond angles and dihedral (torsion) angles were evaluated using the Angles/Torsions measurement functions. Measurements were used to characterize the spatial relationships among amino acid residues, the ligand-binding pocket, and associated structural features.

### Splicing Quantitative (sQTL) Analysis

Splicing quantitative trait locus (sQTL) associations were obtained from GTEx Analysis V10 through the GTEx portal. The four SNPs defining the cocaine use disorder-associated *AACG* haplotype (rs133335, rs7245, rs2284087, and rs5758589) were queried individually, and the resulting association tables were exported as CSV files. Associations annotated to *CYP2D6* were retained, yielding 655 SNP–intron–tissue association records across 46 tissues.

For each association, the GTEx variant identifier, intron identifier, tissue, nominal *p*-value, and normalized effect size (NES) were retained. Because GTEx reports NES relative to the alternative allele, effect sizes were harmonized to the *AACG* haplotype. The NES sign was reversed for rs133335, for which the *AACG* nucleotide (A) is the reference allele, whereas the original NES was retained for rs7245 (A), rs2284087 (C), and rs5758589 (G), for which the *AACG* nucleotide is the alternative allele. Splice-junction associations were compared across SNPs and tissues to identify concordant changes in intron excision phenotypes. These analyses used publicly available GTEx summary statistics; no new sQTL mapping was performed.

## RESULTS

### Long read assemblies

Our phased assemblies for the *CYP2D6* locus from all 19 individuals (38 haploid assemblies) identified 8 mutations comprising 7 haplotypes (Figure 1, Table S4). Five of the 8 mutations are linked in the star 4 (*CYP2D6*4*) haplotype, which includes the splice site disrupting mutation (rs3892097), resulting in a non-functional protein and the poor metabolizer (PM) phenotype (7). The other prevalent haplotype (*CYP2D6*2*) carried two amino acid substitutions (R296C and S486T). Twelve of the fourteen individuals with an *AACG* risk allele carried at least one copy of either *CYP2D6*4* or *CYP2D6*2*. Four of the five individuals that did not carry a copy of the *AACG* risk allele had two fully functional copies identical to the human reference genome. One individual had a normal copy and *9, which has the lysine residue at position 281 deleted.

**Figure 1.**
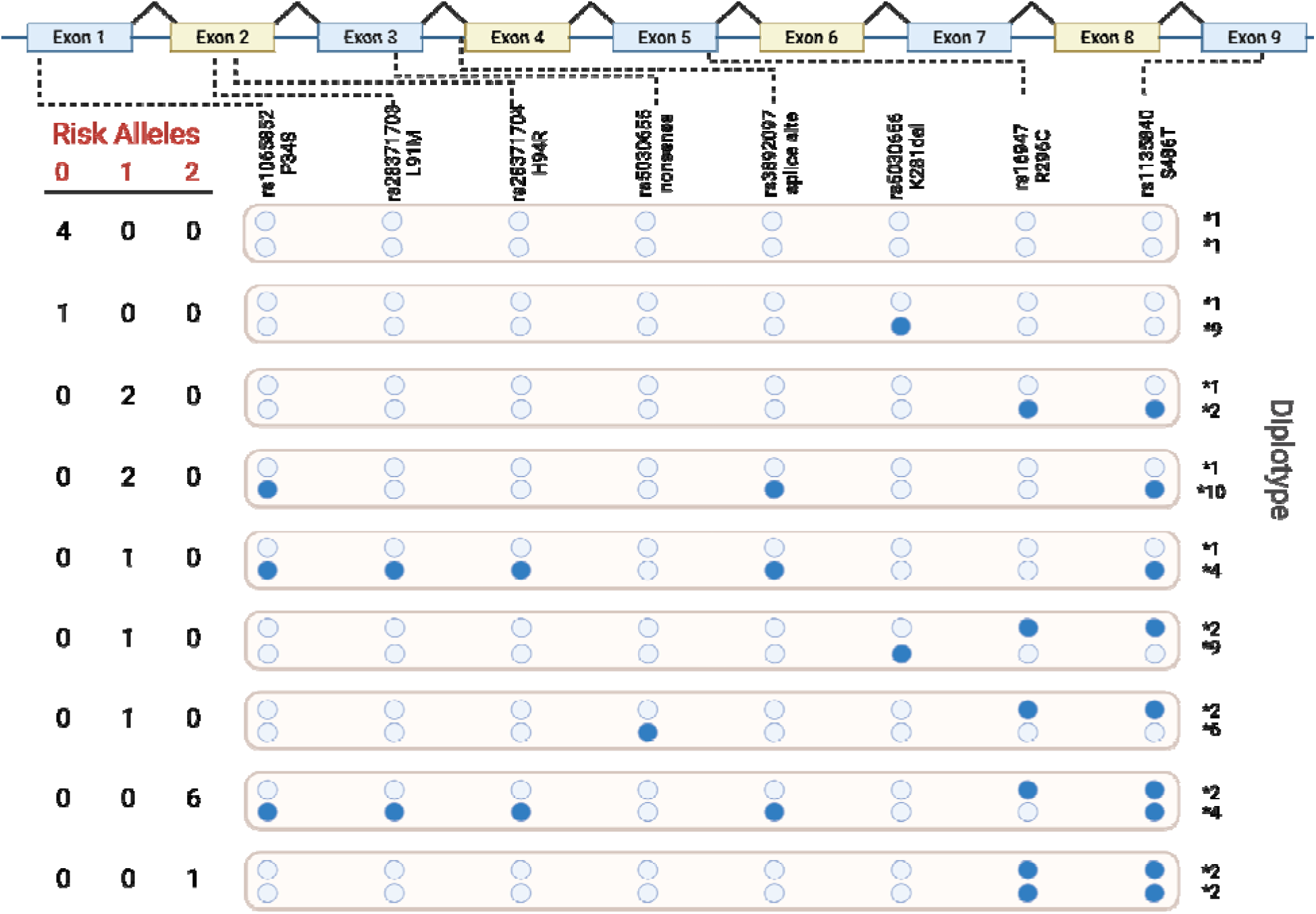
Distribution of phased haplotypes amongst samples that were sequenced with long-read PacBio and their star allele designation from PharmGKB. There are 9 exons in the reference gene model for *CYP2D6* (top). Eight mutations were identified in our panel of 19 individuals. Long reads allowed phasing of the full *CYP2D6* haplotype for each individual. Four of the five individuals with 0 eQTL-risk alleles had two copies of the reference (*\*1*) *CYP2D6* allele. In contrast, none of the individuals with two eQTL-risk alleles carried the reference sequence. Instead, the major haplotype allele in this group carried P34S, L91M, H94R, the splice site change, and S486T (*CYP2D6*4*). A second haplotype found in those with one or two risk alleles carried the R296C and S486T as a linked set (*CYP2D6*2*). Individuals carrying two copies of *CYP2D6*4* are known as poor metabolizers (PM). Those with two copies of *CYP2D6*2* are considered normal metabolizers (NM) but there have been reports of alternative splicing and lower activity for this haplotype (8).

In our previous work we used the mean LogRR intensity values to predict copy number changes at the *CYP2D6* locus. Here we were unable to confirm that these raw intensity values indicated copy number variation given that all 19 individuals carried a *CYP2D6* gene in both alleles. As a potential explanation, we hypothesized the Illumina array itself may explain the observation because two of the SNPs we used for the intensity plot (rs1135835 and rs28371735) overlapped the variant rs1135840 (C/G), which is responsible for the S486T amino acid change. Extraction of the probe sequence revealed that both probes carry the alternate allele “G”, which is present in *\*2*, *\*4*, and *\*10* alleles but not in the *\*1* allele. This is consistent with the probes having reduced affinity given a mismatch in the *\*1* alleles, but having normal binding affinity in the alleles carrying the S486T substitution (Figure 2).

**Figure 2.**
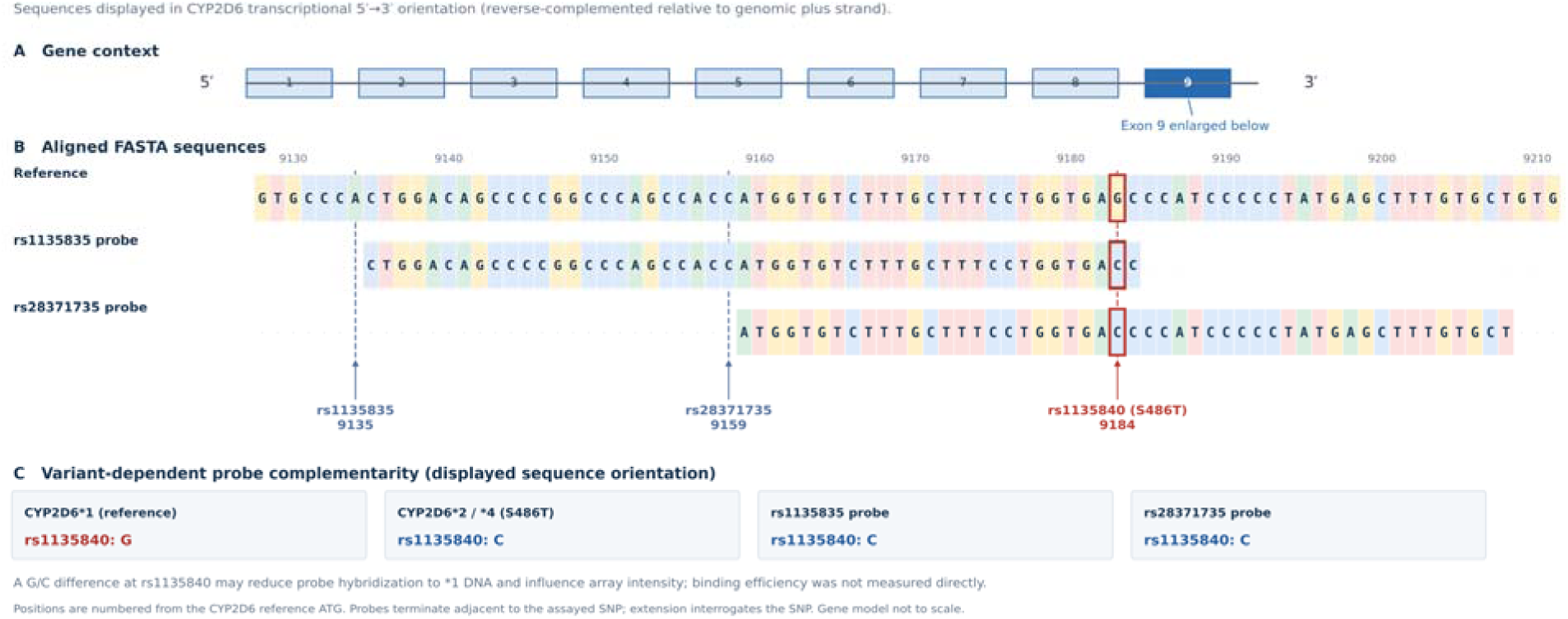
Sequence variation at rs1135840 (S486T) overlaps two Illumina genotyping probes previously used to estimate copy number variation. Alignment of the probe sequences targeting rs1135835 and rs28371735 to the *CYP2D6* reference sequence indicate that both overlap rs1135840, which encodes the S486T amino acid substitution. The probe sequences match the S486T-containing *CYP2D6*2* and *\*4* haplotypes but differ from the *CYP2D6*1* haplotype at this position. This mismatch provides a potential explanation for the reduced array signal intensity observed in individuals carrying *CYP2D6*\*1, independent of underlying copy-number differences.

### Structural predictions for *CYP2D6*2* allele

The *CYP2D6*2* allele is considered to produce a normal metabolizer phenotype but it also has the widest variation in activity depending on the ligand (9). Interestingly, structural comparisons highlight the conformational plasticity of the CYP2D6 protein around residue 486 (Figure 3). In the BACE1 inhibitor–bound structure (5TFT), the S486T substitution lies within 3.8 Å of neighboring residues, suggesting a plausible steric or hydrogen-bonding effect on the substrate channel. In contrast, in the quinidine-bound structure (4WNW), the same residue sits ∼7 Å away, making the structural effect less obvious. These differences support the concept that allelic variants may influence enzyme activity in a substrate-dependent manner, consistent with clinical pharmacogenetic observations (10).

**Figure 3.**
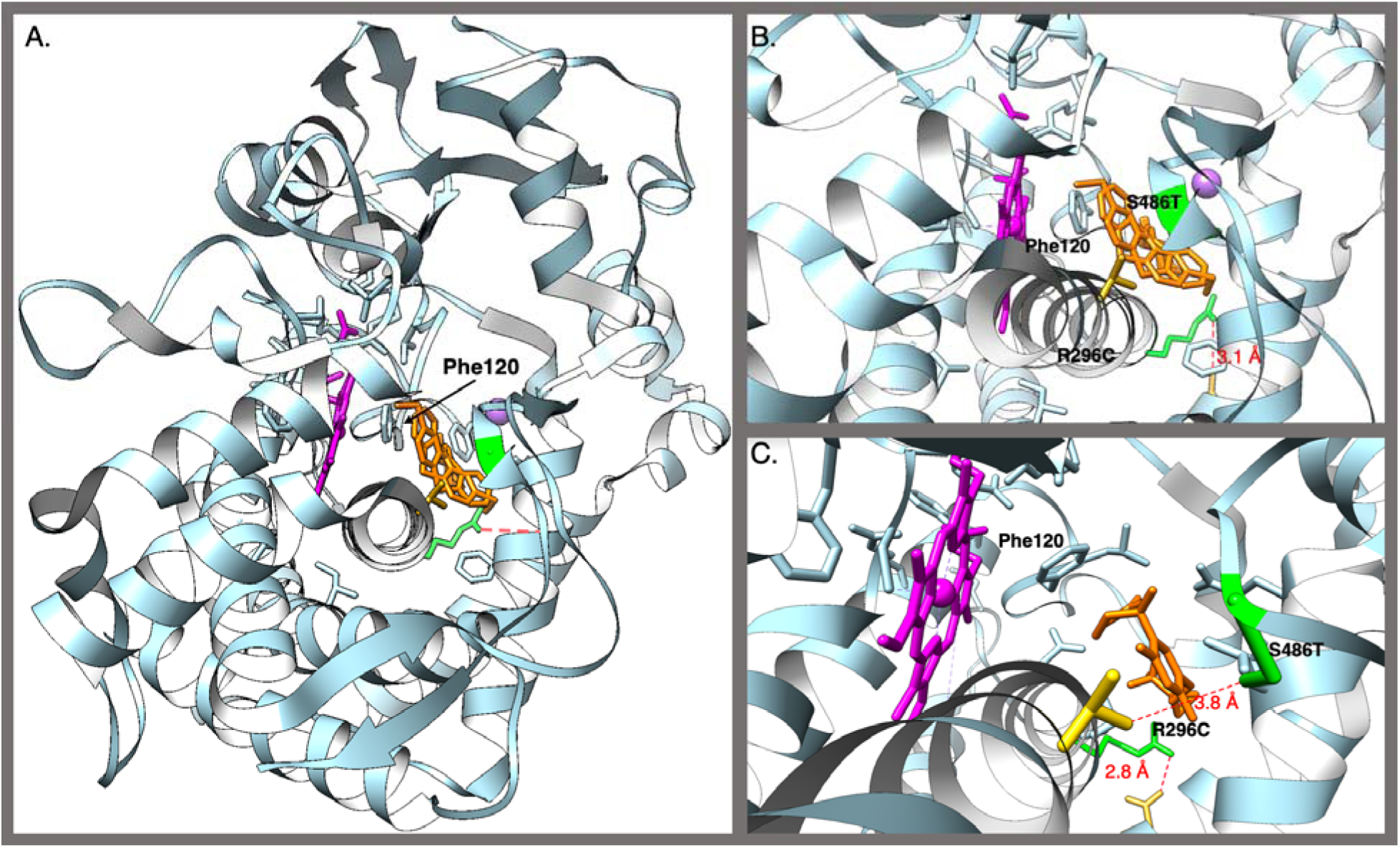
Structural prediction of CYP2D6 and variability of *CYP2D6*2* allele. A. The CYP2D6 structure 4WNW with bound thioridazine (orange) and reactive heme shown in magenta. Phe120 is provided for comparisons across structures. B. Magnified view of the bound ligand showing the two mutations (R296C and S486T) identified in most individuals with the risk haplotype *AACG*. The R296 likely forms a salt bridge with Asp252. C. CYP2D6 PDB structure 5TFT with the BACE1 inhibitor. The salt bridge is retained, but in this structure, the S486T residue is within 5 angstroms of Thr312, suggesting hydrogen bonding and structural alterations from this mutation are likely ligand-dependent whereas the R296C residue is stable with an arginine at this site but absent with the change to cysteine.

### Risk haplotype SNPs are sQTLs across tissues including brain

To investigate whether the cocaine use associated *AACG* haplotype was associated with altered RNA processing, we examined GTEx v10 splicing quantitative trait locus (sQTL) associations for its four constituent SNPs (rs133335, rs7245, rs2284087, and rs5758589). The four variants exhibited highly concordant associations across multiple tissues, including brain regions involved in reward processing. A prominent association involved a short splice junction at chr22:42,142,639–42,142,728 (GRCh38). (Table S5). The associations were observed across multiple tissues, including brain regions relevant to cocaine-induced reward processing. For rs133335, the shorter junction showed particularly strong associations in cerebellum (*p* = 3.3 × 10^-35^, corrected NES = −1.10), cortex (*p* = 2.5 × 10^-34^, corrected NES = −1.00), and cerebellar hemisphere (*p* = 2.4 × 10^-31^, corrected NES = −1.10). Significant associations were also observed in nucleus accumbens (*p* = 9.1 × 10^-22^), caudate (*p* = 5.8 × 10^-30^), putamen (*p* = 1.5 × 10^-21^), and substantia nigra (*p* = 2.9 × 10^-17^).

A second associated junction spanning chr22:42,128,944–42,142,728 crossed the *CYP2D6/CYP2D7* region. Because of the extensive sequence homology between these genes, this signal may reflect ambiguous alignment of RNA-seq reads rather than a genuine splice junction and was therefore not used to infer a specific splicing mechanism. These findings support an association between the *AACG* haplotype and splice-junction usage in multiple tissues. However, the precise transcript consequences and the contribution of the *CYP2D6*4* splice-site variant, rs3892097, remain to be established.

## DISCUSSION

Our previous work identified a four-SNP haplotype within *CYP2D6* strongly associated with cocaine use and altered *CYP2D6* expression in the brain (4). We initially hypothesized this risk haplotype was associated with reduced CYP2D6 enzymatic activity, potentially through structural variation or the presence of nonfunctional alleles. The present study provides a more detailed molecular characterization of this association by resolving *CYP2D6* haplotypes using long-read sequencing. Our results indicate that the risk haplotype is associated with distinct noncanonical *CYP2D6* alleles, particularly *CYP2D6 *2* and *\*4*, rather than tagging gene copy-number variation (CNV). These findings suggest that the relationship between CYP2D6 variation and cocaine use may involve differences in enzyme activity, substrate specificity, and gene regulation.

### Functional diversity of CYP2D6 alleles associated with the risk haplotype

One of the principal findings of this study is that the cocaine use disorder-associated haplotype identifies at least two distinct *CYP2D6* sequence backgrounds with different predicted functional consequences. The *CYP2D6 *4* allele carries four missense substitutions (P34S, L91M, H94R, and S486T) together with the rs3892097 splice-site mutation, whereas the *\*2* allele carries R296C and S486T. Both alleles were observed among individuals carrying the risk haplotype, while four of five individuals lacking the risk haplotype carried two copies of the canonical reference sequence.

These distinctions are important because the established CYP2D6 metabolizer classifications do not necessarily capture the full range of functional differences relevant to endogenous substrates. *CYP2D6 *4* is classified as a non-functional allele, primarily because of its splice-site defect, whereas *\*2* is generally classified as a normal-function allele. However, CYP2D6 activity has been characterized largely through the metabolism of pharmaceutical substrates, and the functional consequences of individual amino acid substitutions may differ across substrates. Furthermore, much of the available pharmacogenetic evidence concerns hepatic metabolism, whereas our previous association analysis identified an effect on *CYP2D6* expression in the brain but not the liver (4).

Consequently, conventional metabolizer classifications may not fully predict the effects of these alleles on endogenous substrate metabolism in neural tissues. Our structural modeling suggests that the missense substitutions associated with the risk haplotype could influence interactions within the CYP2D6 ligand-binding region. Although these predictions require experimental validation, they raise the possibility that alleles classified as having normal activity toward commonly studied pharmaceutical substrates may differ in their metabolism of endogenous compounds. The presence of both *\*2* and *\*4* among risk-haplotype carriers therefore suggests a more complex relationship between genotype and susceptibility than would be expected if the association were attributable exclusively to loss of CYP2D6 function.

### Splice-site disruption and the CYP2D6*4 allele

The *CYP2D6 *4* allele is characterized by the rs3892097 splice-site mutation, which disrupts normal pre-mRNA processing and is associated with markedly reduced or absent CYP2D6 enzymatic activity (11). Our long-read sequencing confirms that this mutation occurs on the same phased haplotype as the P34S, L91M, H94R, and S486T substitutions in the individuals examined here. This provides direct sequence-level evidence linking the cocaine use associated risk haplotype to a pharmacogenomically established non-functional allele.

Analysis of GTEx splice quantitative trait locus (sQTL) data further supports a relationship between variation at this locus and altered RNA processing across multiple tissues, including brain regions involved in reward and reinforcement. The rs3892097 mutation is expected to disrupt canonical splicing and generate transcripts containing premature termination codons, potentially resulting in degradation through nonsense-mediated decay (NMD). Although this mechanism is consistent with the established functional classification of *CYP2D6 *4*, the present study did not directly measure transcript stability or CYP2D6 protein abundance in individuals carrying this allele.

The distinction between altered RNA processing and complete absence of protein is relevant when considering the potential consequences of *CYP2D6 *4* in the brain. A splice-site mutation may substantially reduce the abundance of functional CYP2D6 without necessarily establishing that every transcript or protein product is absent. Additional studies using haplotype-resolved transcript sequencing and protein measurements would help establish the molecular consequences of this allele in neural tissues.

### CYP2D6 sequence variation and evolutionary history

The combination of multiple missense substitutions and a splice-site mutation on the *CYP2D6 *4* haplotype also raises questions about the evolutionary history of this allele. The coding substitutions may have arisen before the splice-site mutation and could have influenced substrate interactions prior to the loss of normal splicing. Alternatively, these variants may have accumulated through other evolutionary processes, including genetic drift, linkage, or selection acting on neighboring variants.

The relative order in which these mutations arose cannot be determined from the present data alone. Nevertheless, the persistence of a haplotype carrying several amino acid substitutions together with a loss-of-function splice-site mutation suggests that the evolutionary history of *CYP2D6* may be more complex than a simple accumulation of deleterious changes. Comparisons of phased haplotypes across diverse human populations, together with ancestral sequence reconstruction and tests of selection, could help determine whether these coding substitutions preceded the splice-site mutation and whether any were subject to positive selection. An important unresolved question is whether the amino acid substitutions associated with *CYP2D6 *2* and *\*4* influence the metabolism of endogenous substrates. If so, these variants may have functional consequences that are not apparent from conventional pharmaceutical-based activity classifications.

### Implications for dopamine metabolism and cocaine susceptibility

The potential relevance of these findings to cocaine use extends beyond the established role of CYP2D6 in drug metabolism. CYP2D6 has been implicated in the metabolism of several endogenous compounds, including pathways related to dopamine and trace amines (12–15). In our previous study, the four-SNP risk haplotype was associated with altered *CYP2D6* expression in the brain, suggesting that its functional consequences may differ between neural and hepatic tissues (4).

Cocaine produces its reinforcing effects in part by inhibiting dopamine reuptake and increasing extracellular dopamine concentrations in reward-related neural circuits. Genetic variation that alters CYP2D6-mediated metabolism of endogenous substrates could therefore influence dopaminergic signaling or the response to cocaine. The presence of a non-functional *\*4* allele among risk-haplotype carriers is consistent with a potential reduction in CYP2D6-dependent metabolic activity. In contrast, the *\*2* allele may retain substantial enzymatic activity while exhibiting substrate-dependent differences that are not captured by standard metabolizer classifications.

This functional heterogeneity may be particularly relevant to our previous observation that the association between the *CYP2D6* haplotype and cocaine use was dependent on the haplo-diplotype configuration. The combination of alleles inherited by an individual may be more informative than the presence or absence of a single risk haplotype, particularly if different CYP2D6 proteins vary in their interactions with endogenous substrates. However, the present study does not directly establish that the identified alleles alter dopamine concentrations, dopamine metabolite ratios, or cocaine-induced neuronal responses. Future studies should therefore examine the effects of the identified haplotypes on CYP2D6 protein abundance, substrate-specific enzymatic activity, and dopamine-related metabolic pathways. Such experiments would help determine whether the association with cocaine use reflects reduced enzyme activity, altered substrate specificity, regulatory differences, or a combination of these mechanisms.

### Interpretation of SNP-array intensity and copy-number variation

An additional finding of this study concerns the interpretation of genotyping-array signal intensity at the structurally complex *CYP2D6* locus. Our initial analyses identified differences in raw array intensity that correlated with the number of risk-haplotype copies, suggesting that the associated haplotype might tag underlying CNV. However, analysis of long-read assemblies and targeted sequencing did not support a consistent relationship between the risk haplotype and *CYP2D6* gene copy number. Instead, examination of the Illumina probe sequences revealed that neighboring polymorphisms could produce mismatches within probe-binding regions, potentially affecting hybridization efficiency and measured signal intensity. In particular, the rs1135840 variant overlaps the binding regions of probes targeting rs1135835 and rs28371735. The resulting sequence differences provide a plausible explanation for the apparent copy-number signal without requiring a corresponding change in gene dosage.

These findings illustrate an important limitation of interpreting SNP-array intensity as a proxy for CNV at highly polymorphic loci. Sequence differences within probe-binding regions may produce haplotype-dependent intensity changes that resemble structural variation (16). Accordingly, apparent CNV associations at complex pharmacogenomic loci should be interpreted cautiously and, where possible, evaluated using haplotype-resolved sequencing or independent copy-number assays.

### Limitations and conclusions

We note several limitations from our study. Targeted long-read sequencing was performed in 19 individuals, providing detailed molecular characterization but limiting the ability to estimate the population frequencies of specific *CYP2D6* alleles or establish their independent contributions to cocaine use risk. In addition, the predicted effects of the missense substitutions on ligand binding were derived from structural modeling rather than direct enzymatic measurements. Although the *CYP2D6 *4* splice-site mutation has an established loss-of-function classification, its molecular consequences were not directly assessed in brain tissue from the sequenced individuals.

Despite these limitations, our findings provide an important molecular extension of the previously reported association between a four-SNP *CYP2D6* haplotype and cocaine use (4). Long-read sequencing demonstrates that the associated haplotype marks distinct *CYP2D6* alleles, including *\*2* and *\*4*, with different established or predicted functional properties. These results also indicate that the previously observed SNP-array intensity differences are more consistent with probe-sequence effects than with effects consistent with underlying CNV. Together, these findings support further investigation of CYP2D6-mediated endogenous metabolism as a potential contributor to cocaine susceptibility and highlight the importance of resolving complete haplotypes when interpreting genetic associations at complex pharmacogenomic loci.

## INSTITUTIONAL REVIEW BOARD

The study was reviewed and approved by the Rutgers Institutional Review Board.

## DATA AVAILABILITY STATEMENT

Sequences produced for this study are provided as FASTA files in supplementary data and in supplementary tables.

## ACKNOWLEDGEMENTS AND FUNDINGS

Research reported in this publication was supported by the National Institute on Drug Abuse (NIDA) of the National Institutes of Health under Award Numbers R01DA050495, as well as the Laboratory Directed Research & Development Program of Oak Ridge National Laboratory, managed by UT-Battelle, for the US Department of Energy (DOE). Also supported by the New Jersey Commission on Cancer Research under grant number COCR26PPR013. Biospecimens were genotyped under NIDA Contract N01DA187789. The content is solely the responsibility of the authors and does not necessarily represent the official views of NIDA, the National Institutes of Health, the US DOE, or the New Jersey Commission on Cancer Research.

## AUTHOR CONTRIBUTIONS

MG – Conceptualization, Methodology, Validation, Formal Analysis, Investigation, Writing the Original Draft, Review and Editing, Visualization, Funding Acquisition. AWB – Conceptualization, Review and Editing. DMR – Conceptualization, Review and Editing, Data Curaton. SHW – Conceptualization, Review and Editing, Funding Acquisition.

